# Aneurysmal versus “Benign” Perimesencephalic Subarachnoid Hemorrhage

**DOI:** 10.1101/2023.07.17.23292803

**Authors:** Anas Alrohimi, Mark A Davison, Abhi Pandhi, Mohammad A Abdulrazzak, Daniel Wadden, Mark Bain, Nina Moore, M. Shazam Hussain, Gabor Toth

## Abstract

**Introduction:** The rate of underlying ruptured aneurysm, complications, and outcomes in “benign” perimesencephalic subarachnoid hemorrhage (PMSAH) are not well known and underreported.

**Methods:** Retrospective analysis of patients with PMSAH from a large tertiary care center (2007-2023). Clinical and imaging data were studied.

**Results:** Eighty-one patients were included with mean age of 55.5 ± 10.3 years. Median (IQR) Hunt and Hess grade was 2 (1-2), and modified Fisher grade was 3 (1-3). An underlying ruptured aneurysm was diagnosed in 5 patients (6.2%). The most common complication was vasospasm in 25 patients (31%) with a significantly higher rate of symptomatic vasospasm among patients with underlying aneurysm (40% vs 2.6%; P=0.01). The median (IQR) time of vasospasm detection was 7 (8-9) days. The majority of vasospasm cases (84%) were asymptomatic, which was not associated with poor outcomes (OR= 0.95, [0.22–4.1], *P*=0.9) compared to the asymptomatic vasospasm (OR= 8.6, [1.06-69.88], *P*= 0.04). Hydrocephalus occurred in 10% of patients within one day, at higher rate in the aneurysmal group (40% vs 8%; P=0.07). A total of 88% of patients had a favorable functional outcome (mRS 0-2) at discharge, but at a significantly higher rate in non-aneurysmal patients (91% vs 40%; P=0.01). An underlying aneurysm and hydrocephalus were associated with poor functional status (OR= 14.7, [2.1–104], *P*=0.007, and OR= 22.6, [4.2–123.5], *P* <0.001), respectively.

**Conclusion:** “Benign” PMSAH pattern was associated with a ruptured aneurysm in 6.2% of patients, which highlights the critical value of conventional cerebral angiogram in the workup. Expectedly, an underlying aneurysm was associated with higher rates of symptomatic vasospasm, hydrocephalus, and lower rates of good clinical outcome. Aneurysm and hydrocephalus were independently associated with poorer outcomes, however, these were detected very early during hospitalization. Vasospasm alone was not associated with poor outcomes. Our findings suggest that non-aneurysmal PMSAH patients can safely be managed with less strict monitoring and a shorter hospital stay.

## INTRODUCTION

Non-traumatic subarachnoid hemorrhage is a prevalent and often devastating pathology which accounts for approximately 5% of all strokes in the United States.^(1)^ Potential etiologies for a spontaneous subarachnoid bleed are many, and while an intracranial aneurysm is frequently the culprit, an underlying etiology is not always evident in up to 15-20% of cases.^(2, 3)^ Benign perimesencephalic subarachnoid hemorrhage (PMSAH) was first described in 1985 as a subset of patients with SAH that was identified with a pattern of hemorrhage centered anterior to the midbrain without frank extension to the anterior interhemispheric fissure, lateral sylvian fissure intraparenchymal or intraventricular, without an identifiable vascular lesion, and a benign clinical course.^(3–5)^

Diagnostic investigation of spontaneous subarachnoid hemorrhage often begins with computed tomography angiography (CTA), however digital subtraction angiography (DSA) remains the gold standard for identifying intracranial vascular pathology. Given that CTA technology continues to advance, some reports suggest that providers may be able to forego DSA in patients with PMSAH following a negative CTA.^(6)^ While the yield of DSA in the PMSAH population following a negative CTA is low, the consequences of missing an occult intracranial aneurysm could be devastating.

The term “benign” is often associated, because typically no source of bleeding is identified on high-resolution vascular imaging and recovery is often uncomplicated. However, PMSAH can be secondary to ruptured vertebrobasilar aneurysms in 5-7% of patients and outcomes in these patients is underreported.^(5, 7)^ Moreover, while relatively infrequent, patients with PMSAH have been known to suffer from symptomatic hydrocephalus or vasospasm, which may confer a significant morbidity to the patient.^(8)^ Therefore, it is essential to consider a large patient cohort while assessing the incidence of a relatively infrequent event and study clinical outcomes. We conducted a retrospective analysis of patients with PMSAH in a large single center to determine the rate of underlying ruptured aneurysm, associated complications, its timeline, outcomes, and report the diagnostic yield of DSA.

## METHODS

### Study design

The current study is a retrospective analysis of PMSAH patients treated at a large single-center institution from 2007-2023. Eligible patients were identified via ENCORE radiology angiogram reports database. Specifically, reports including either the keywords (perimesencephalic, and perimesencephalic subarachnoid hemorrhage) were selected. The non-contrast CT brain scan of all identified patients were assessed to confirm an accurate radiographic diagnosis of PMSAH. CT angiogram of the brain were reviewed as well. Patients with both a diagnosis of PMSAH, available medical records and images for review were included. Age, sex, vascular risk factors (hypertension, smoking, and family history of intracranial aneurysms), clinical presentation, Hunt and Hess scale, modified Fisher grade, and rate of underlying ruptured aneurysm were collected. Vasospasm based on clinical, sonographic and angiographic diagnosis, hydrocephalus based on clinical and radiographic diagnosis, the number of patients required external ventricular drain (EVD) and discharged modified Rankin scale (mRS) were gathered. Data was analyzed for all patients, as well as the aneurysmal and non-aneurysmal groups separately for comparison. Institutional Review Boards approval was obtained.

### Imaging Procedures and Analysis

All patients enrolled in the analysis had a non-contrast head computed tomography (CT), CT angiography, and digital subtraction angiography (DSA) at baseline. In the event of clinical deterioration, CT scans were repeated. In the event of clinical deterioration with sonographic diagnosis of vasospasm, DSA was repeated. All images were independently analyzed by three endovascular surgical neuroradiology fellows (AA, AP and MAA), and any disagreement was resolved by consensus.

### Outcomes

The primary endpoint was good functional status at discharge, which was defined as a Modified Rankin Score (mRS) of 0-2. The incidence of an underlying ruptured aneurysm through DSA, and whether it is present in CTA, was reported. Additionally, clinical complications including the rate of hydrocephalus and vasospasm were documented.

### Statistical analysis

Statistical analyses were performed using the Statistical Package for Social Sciences version 28.0.0 (IBM SPSS Statistics Inc, 2015, Armonk, NY, USA). Baseline characteristics were reported by mean and standard deviations (SD) for continuous variables with normal distribution, median and interquartile ranges (IQRs) for continuous variables with skewed distribution, frequency and percentage for categorical variables. Differences between groups were assessed using independent t-tests for parametric data, and Mann-Whitney-U tests for non-parametric data. The distribution of outcomes was assessed with Pearson’s χ^2^ test or Fisher’s exact test. Univariable and Multivariable (if required) logistic regression models were used to determine factors that are independently associated with ruptured aneurysm and outcomes. Functional outcome at discharge was dichotomized as independent (mRS score 0-2) and dependent (mRS score 3-6).

## RESULTS

### Patient characteristics

A total of 86 patients identified between 2007 and 2023 were assessed for eligibility. Of these, 81 patients met the criteria and were included in the analysis. An underlying ruptured aneurysm was found in 5 patients (6.2%); four of which were in the vertebrobasilar system and one in the posterior communicating artery. Baseline characteristics of patients with and without underlying aneurysm is summarized in **Table 1**. Mean patient age was 55.5 ± 10.3 years and 62% were male. Among vascular risk factors, hypertension and smoking were reported in 55% and 35% of all patients, respectively. Only one patient reported family history of ruptured intracranial aneurysm, in the non-aneurysmal PMSAH group. With regards to the presenting symptoms, the most common symptom was thunderclap headache in 89% of patients. Cranial neuropathy and extremity weakness were reported in 6% and 4%, respectively. Median (IQR) of Glasgow coma scale was 15 (15-15). Median (IQR) of Hunt and Hess scale was 2 (1-2), and modified Fisher grade was 3 (1-3).

**Table 1.** Baseline patients’ characteristics.

|  | Patients with underlying ruptured aneurysm (n=5) | Patients with negative angiogram (n=76) | *P value* |
| --- | --- | --- | --- |
| Age, mean ± SD | 56 ± 10.2 years | 55.5 ± 10.3 years | 0.9 |
| Male, n (%) | 4 (80) | 46 (60.5) | 0.6 |
| Hypertension, n (%) | 5 (100) | 40 (52.5) | 0.06 |
| Smoking, n (%) | 4 (80) | 24 (31.5) | 0.04 |
| Family history of ruptured aneurysm, n (%) | 0 (0) | 1 (1.3) | 1.0 |
| Headache, n (%) | 5 (100) | 67 (88) | 1.0 |
| Cranial neuropathy, n (%) | 0 (0) | 2 (2.5) | 0.17 |
| Focal deficit, n (%) | 1 (20) | 5 (6.5) | 1.0 |
| GCS, median (IQR) | 15 (14-15) | 15 (15-15) | 0.07 |
| Hunt and Hess scale, median (IQR) | 3 (1.5-3) | 2 (1-2) | 0.04 |
| modified Fisher grade, median (IQR) | 4 (1-4) | 3 (1-3) | 0.01 |

### PMSAH workup and patient outcomes

An underlying ruptured aneurysm was found in 5 patients (6.2%); three of which (60%) were detected in DSA (two on initial DSA and one on follow-up DSA), and the median (IQR) time of aneurysmal detection was 1 (1-2) day. Smoking (*P*=0.04), Hunt and Hess scale (*P*=0.04), and modified Fisher grade (*P*=0.01) were significantly higher in patients with aneurysmal PMSAH. A total of 71 (88%) patients in this cohort had a repeat diagnostic angiogram. The most common complication in the whole cohort was vasospasm in 25 (31%) patients; the majority of which (84%) were asymptomatic. Among the aneurysmal PMSAH group, symptomatic vasospasm was detected in 2 out of 5 patients (40%) which was significantly higher than the non-aneurysmal PMSAH group (2 out of 76 patients, 2.6%; *P*= 0.01). All 4 patients with symptomatic vasospasm received intra-arterial vasodilation therapy, and two received additional angioplasty. The median (IQR) time of all vasospasm detection was 7 (8-9) days, and asymptomatic vasospasm was not associated with unfavourable outcomes (OR= 0.95, [0.22–4.1], *P*= 0.9). However, symptomatic vasospasm was associated with worse outcomes (OR= 8.6, [1.06-69.88], *P*= 0.04). Acute hydrocephalus occurred in overall 10% of patients, at a numerically higher rate in the aneurysmal group, however, the difference was not statistically significant (40% vs 8%; *P*= 0.07). All hydrocephalus cases were symptomatic, and occurred within 1 day, but at a significantly higher rate in the aneurysmal group (2 out of 5 patients, 40%). In the entire cohort, median (IQR) of mRS at discharge was 0 (0-1), and a total of 71 (88%) patients had a favorable functional outcome (mRS 0-2) at discharge. The median (IQR) mRS at discharge of patients with aneurysmal PMSAH pattern was 3 (1-4) which was not significantly higher than the non-aneurysmal group 0 (0-1); *P*=0.07. However, the distribution of mRS between the two groups was significantly different, *P*= 0.015. The favorable functional outcome (mRS 0-2) at discharge in the aneurysmal PMSAH pattern group (40%) was significantly lower than the non-aneurysmal group (91%); *P*=0.01. In our analysis, an underlying ruptured aneurysm and acute hydrocephalus were associated with poor functional status (OR= 14.7, [2.1–104], P=0.007; Figure 1), and OR= 22.6, [4.2–123.5], P <0.001), respectively.

**Figure 1.**
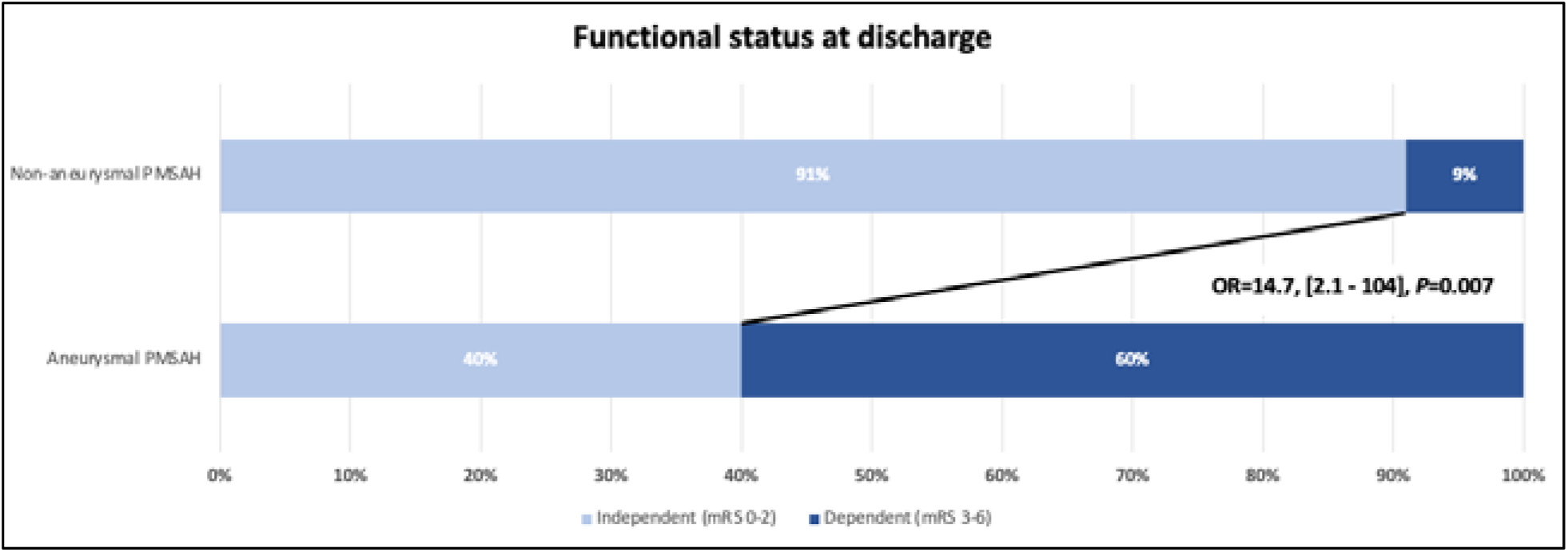
Functional status at discharge of patients with aneurysmal and non-aneurysmal source of PMSAH.

## DISCUSSION

In this retrospective analysis of 81 patients with PMSAH, the rate of underlying ruptured aneurysm through catheter angiography was 6.2%. All aneurysms were found in the posterior circulation. The presence of an underlying ruptured aneurysm was associated with worse clinical outcomes, and higher rates of symptomatic vasospasm and hydrocephalus. The low but non-negligible rate and serious clinical consequences highlight the sustained critical value of conventional cerebral angiogram in the diagnostic workup of PMSAH. On the other hand, the low rates of symptomatic hydrocephalus and vasospasm, and early appearance of acute hydrocephalus in the non-aneurysmal PMSAH group provide clinical evidence for less strict clinical monitoring, earlier discharge and decreased length of hospitalization for these patients.

The current study is one of the largest published cohorts of patients with PMSAH to date, and is the largest series which has included patients within the last decade. Our extensive literature review resulted in 16 studies including over 650 patients with PMSAH pattern on conventional CT brain and a negative CTA. Diagnostic cerebral angiography was found to be positive for an occult aneurysm in approximately 3.2 – 3.5% of patients (**Table 3**). While our finding of a 6.2% aneurysm identification rate was slightly higher, these findings are clinically significant when considering the devastating consequences of missing an occult ruptured aneurysm: re-rupture rates are known to reach 50% in 6 months, with the highest risk occurring in the first 48 hours.^(2, 9)^ Interestingly, the vast majority (approximately 81% in the literature and 60% in our cohort) of aneurysms identified on DSA who had negative CTA were in the vertebrobasilar circulation, which could be related to increased bone artifact from the posterior fossa.

**Table 2.** Patients’ outcomes.

|  | Patients with underlying ruptured aneurysm (n=5) | Patients with negative angiogram (n=76) | *P value* |
| --- | --- | --- | --- |
| Modified Rankin Scale, median (IQR) | 3 (1-4) | 0 (0-1) | 0.07 |
| Favorable functional outcome (mRS 0-2) at discharge | 2 (40) | 69 (91) | 0.01 |
| Symptomatic vasospasm, n (%) | 2 (40) | 2 (2.6) | 0.01 |
| Symptomatic hydrocephalus, n (%) | 2 (40) | 6 (8) | 0.07 |

**Table 3.** Diagnostic yield of digital subtraction angiography in the PMSAH population; the current study and summary of the literature.

| Reference | Data Collection Range | PMSAH Patients | Patients with underlying vascular pathology on angiogram | Aneurysms identified |
| --- | --- | --- | --- | --- |
| Alrohimi et al., 2023 | 2007 - 2023 | 81 | 5 (6.2%) | Vertebrobasilar (4)<br>Posterior Communicating Artery (1) |
| Catapano et al. <sup>(12)</sup> | 2014 - 2018 | 49 | 0 (0%) | No lesions found |
| Mortimer et al. <sup>(6)</sup> | 2006 - 2014 | 72 | 1 (1.4%) | Vertebral Artery Dissection (1) |
| Kalra et al. <sup>(13)</sup> | 2008 - 2014 | 18 | 0 (0%) | No lesions found |
| Zhong et al. <sup>(14)</sup> | 2009 - 2012 | 49 | 0 (0%) | No lesions found |
| Delgado - Almandoz et al. <sup>(15)</sup> | 2007 - 2011 | 29 | 1 (3.4%) | Left superior cerebellar (1) |
| Moscovici et al. <sup>(16)</sup> | 2005 - 2011 | 25 | 0 (0%) | No lesions found |
| Cruz et al. <sup>(17)</sup> | 2006 - 2010 | 41 | 1 (2.4%) | ICA aneurysm (1) |
| Agid et al. <sup>(18)</sup> | 2005 - 2009 | 93 | 0 (0%) | No lesions found |
| Little et al. <sup>(19)</sup> | 2003 - 2006 | 16 | 1 (6.3%) | Anterior Inferior Cerebellar (1) |
| Westerlaan et al. <sup>(20)</sup> | 2003 - 2006 | 30 | 0 (0%) | No lesions found |
| Kershenovich et al. <sup>(21)</sup> | 2001 - 2005 | 30 | 0 (0%) | No lesions found |
| Alén et al. <sup>(22)</sup> | 1990 - 2000 | 45 | 4 (8.9%) | Vertebrobasilar (4) |
| van Dijk et al. <sup>(23)</sup> | < 2001 | Observer 1: 26<br>Observer 2: 38 | Observer 1: 7 (26.9%)<br>Observer 2: 9 (23.7%) | Observer 1: ICA (2),<br>Vertebrobasilar (5)<br>Observer 2: ICA (2),<br>Vertebrobasilar (7) |
| Velthuis et al. <sup>(24)</sup> | 1994 - 1997 | 15 | 0 (0%) | No lesions found |
| Rinkel et al. <sup>(3)</sup> | 1983 - 1989 | Observer A: 20<br>Observer B: 16 | Observer A: 1 (5%)<br>Observer B: 1 (6.3) | Observer A: Vertebrobasilar (1)<br>Observer B: Vertebrobasilar (1) |

In contrast to classic aneurysmal SAH, the majority of patients with benign PMSAH pattern were male (62%) with a mean age of 55.5 years. This demographic finding in our population is consistent with prior reports.^(5, 10)^ The higher rate of hypertension and smoking, though the former was not significant, in the aneurysmal PMSAH group is not surprising, and confirms the relevance of these vascular risk factors in aneurysm formation and rupture.^(11)^ Compared to classic aneurysmal SAH, patients with PMSAH tend to present with milder symptoms with lower Hunt and Hess scale and modified Fisher grade, which was also demonstrated in our cohort.^(5)^ This finding suggests that an underlying ruptured aneurysm should be considered and searched for when interpreting vascular imaging of patients with a more severe clinical profile, and one should not hesitate to obtain a conventional angiogram for these patients.

Complications such as vasospasm and hydrocephalus were reported previously to be infrequent after PMSAH compared to aneurysmal SAH.^(5)^ While radiographic vasospasm was observed in overall 31% in our cohort, only 4 patients of the cohort (4.9%) were symptomatic; 2 (40%) in the aneurysmal and 2 (2.6%) in the non-aneurysmal group. The median time of all vasospasm detection was 7 (8-9) days. Our results confirm that symptomatic vasospasm in non-aneurysmal PMSAH patients is a rare phenomenon, only described previously in case reports or small patient series. However, vasospasm, even if present, was not associated with worse outcome in our cohort. Contrary to the historical belief about benign PMSAH, acute hydrocephalus can occur in this group as was seen in 8% of our non-aneurysmal PMSAH population, which was still much lower than the aneurysmal group (40%). All cases occurred within 1 day, suggesting the hydrocephalus risk is highest soon after the index event, which decreases the need for strict prolonged hospital monitoring for these patients. Unlike vasospasm in this cohort, which was clinically silent in the majority, acute hydrocephalus events were associated with worse functional outcomes. Similar to previous reports, favorable functional outcomes (mRS 0-2) at time of discharge were observed in the majority of patients with non-aneurysmal (91%) which was significantly higher than the aneurysmal group (91% vs. 40%; *P*= 0.01).

### Limitations

While the results of the current study are compelling, they must be interpreted within the context of the study limitations. First, our study is subject to all the limitations inherent to a retrospective analysis comprised of subjects from a single center. In addition, while the majority of our cohort (88%) had a repeat follow up DSA, under reporting underlying aneurysms in the remaining 12% who did not have a follow up DSA is a possibility. However, even with the above limitations, this is one of the largest studies available on PMSAH patients providing valuable information for clinical decision making.

## CONCLUSIONS

In our cohort, “benign” PMSAH pattern was associated with a ruptured aneurysm in 6.2% of patients, and we advocate for conventional cerebral angiogram to remain part of the diagnostic workup. Expectedly, acute hydrocephalus was seen very early during the hospital course, mostly with an underlying aneurysm, and the presence of these two were associated with poorer outcomes. Vasospasm alone was not associated with unfavorable outcomes in patients with PMSAH. Our findings support less strict monitoring, and shorter hospitalization of patients with PMSAH without an underlying ruptured aneurysm or acute hydrocephalus.

## Data Availability

The data referred to in the manuscript is available

## ACKNOWLEDGMENTS

We thank and acknowledge Vickie Keller, RT(R), Thomas Frank, RT(R), Elizabeth Beno, RT(R), and William (Bill) Tajgiszer, RT(R) in imaging institute at Cleveland Clinic for their assistance in retrieving images from the imaging archives.

---

STROBE Statement—checklist of items that should be included in reports of observational studies

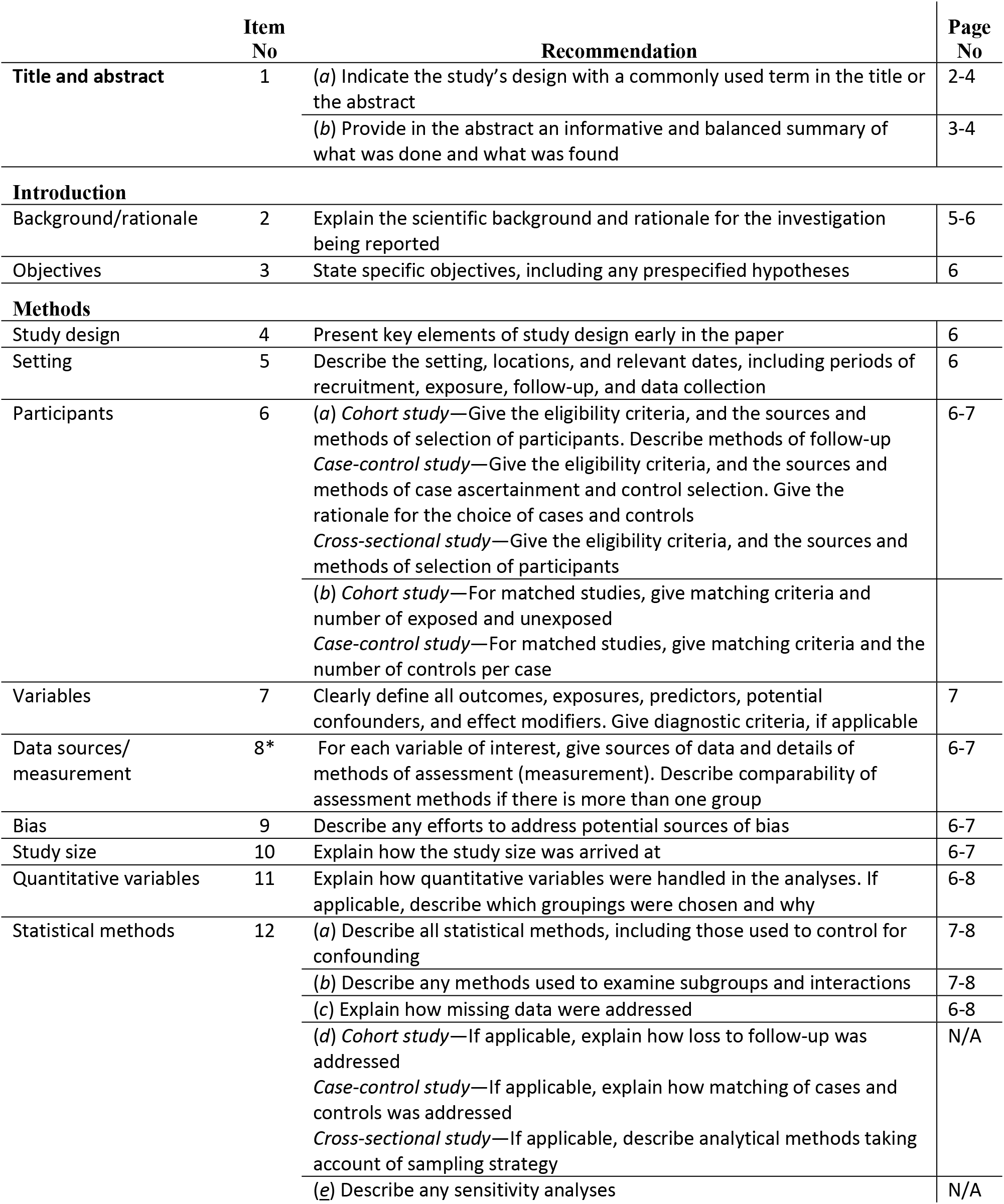
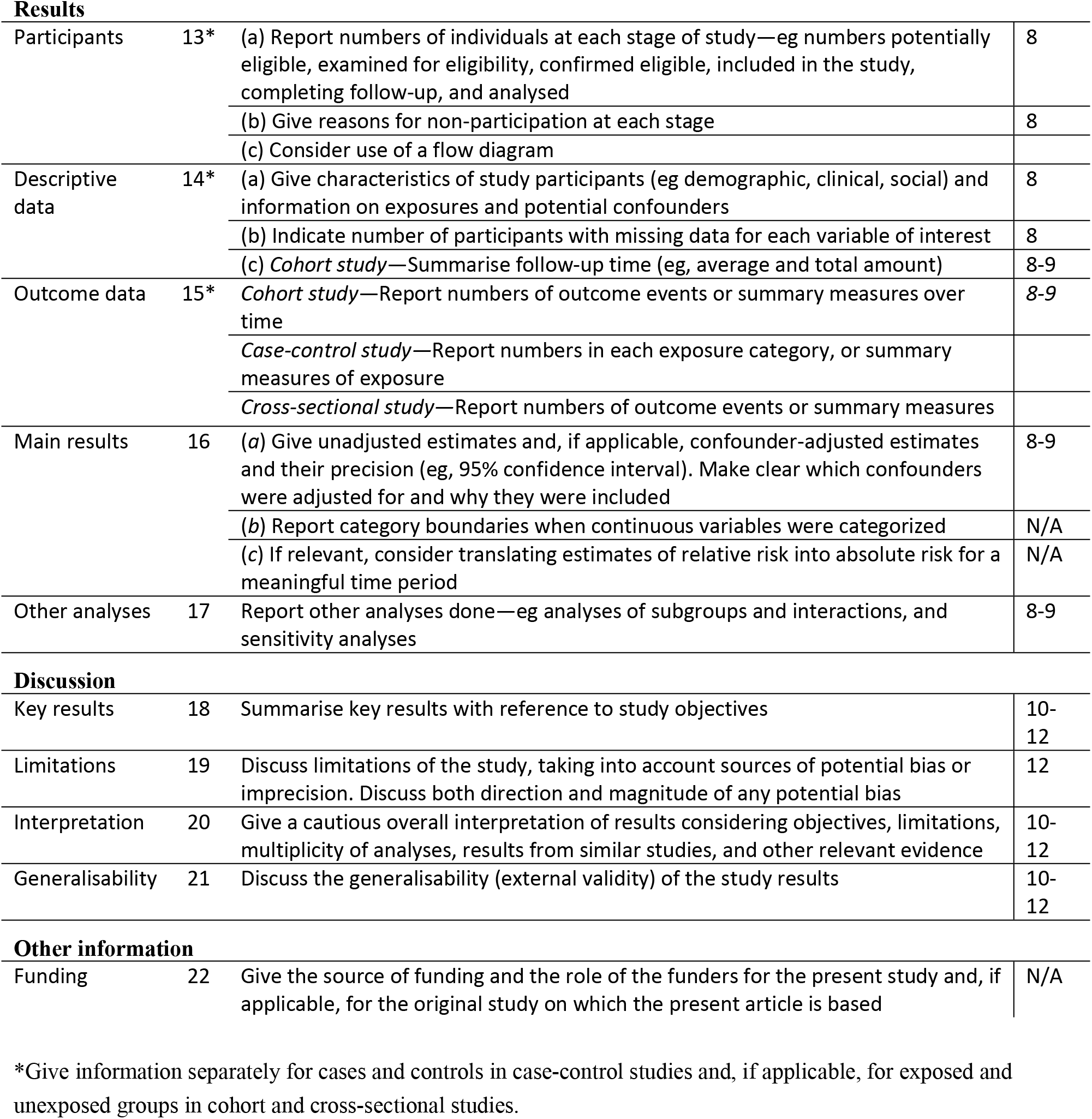

|  | Item No | Recommendation | Page No |
| --- | --- | --- | --- |
| Title and abstract | 1 | (a) Indicate the study’s design with a commonly used term in the title or the abstract | 2-4 |
|  |  | (b) Provide in the abstract an informative and balanced summary of what was done and what was found | 3-4 |
| Introduction |  |  |  |
| Background/rationale | 2 | Explain the scientific background and rationale for the investigation being reported | 5-6 |
| Objectives | 3 | State specific objectives, including any prespecified hypotheses | 6 |
| Methods |  |  |  |
| Study design | 4 | Present key elements of study design early in the paper | 6 |
| Setting | 5 | Describe the setting, locations, and relevant dates, including periods of recruitment, exposure, follow-up, and data collection | 6 |
| Participants | 6 | (a) Cohort study—Give the eligibility criteria, and the sources and methods of selection of participants. Describe methods of follow-up<br>Case-control study—Give the eligibility criteria, and the sources and methods of case ascertainment and control selection. Give the rationale for the choice of cases and controls<br>Cross-sectional study—Give the eligibility criteria, and the sources and methods of selection of participants | 6-7 |
|  |  | (b) Cohort study—For matched studies, give matching criteria and number of exposed and unexposed<br>Case-control study—For matched studies, give matching criteria and the number of controls per case |  |
| Variables | 7 | Clearly define all outcomes, exposures, predictors, potential confounders, and effect modifiers. Give diagnostic criteria, if applicable | 7 |
| Data sources/measurement | 8* | For each variable of interest, give sources of data and details of methods of assessment (measurement). Describe comparability of assessment methods if there is more than one group | 6-7 |
| Bias | 9 | Describe any efforts to address potential sources of bias | 6-7 |
| Study size | 10 | Explain how the study size was arrived at | 6-7 |
| Quantitative variables | 11 | Explain how quantitative variables were handled in the analyses. If applicable, describe which groupings were chosen and why | 6-8 |
| Statistical methods | 12 | (a) Describe all statistical methods, including those used to control for confounding | 7-8 |
|  |  | (b) Describe any methods used to examine subgroups and interactions | 7-8 |
|  |  | (c) Explain how missing data were addressed | 6-8 |
|  |  | (d) Cohort study—If applicable, explain how loss to follow-up was addressed<br>Case-control study—If applicable, explain how matching of cases and controls was addressed<br>Cross-sectional study—If applicable, describe analytical methods taking account of sampling strategy | N/A |
|  |  | (e) Describe any sensitivity analyses | N/A |
| **Results** |  |  |  |
| Participants | 13* | (a) Report numbers of individuals at each stage of study—eg numbers potentially eligible, examined for eligibility, confirmed eligible, included in the study, completing follow-up, and analysed | 8 |
|  |  | (b) Give reasons for non-participation at each stage | 8 |
|  |  | (c) Consider use of a flow diagram |  |
| Descriptive data | 14* | (a) Give characteristics of study participants (eg demographic, clinical, social) and information on exposures and potential confounders | 8 |
|  |  | (b) Indicate number of participants with missing data for each variable of interest | 8 |
|  |  | (c) *Cohort study*—Summarise follow-up time (eg, average and total amount) | 8-9 |
| Outcome data | 15* | *Cohort study*—Report numbers of outcome events or summary measures over time | 8-9 |
|  |  | *Case-control study*—Report numbers in each exposure category, or summary measures of exposure |  |
|  |  | *Cross-sectional study*—Report numbers of outcome events or summary measures |  |
| Main results | 16 | (a) Give unadjusted estimates and, if applicable, confounder-adjusted estimates and their precision (eg, 95% confidence interval). Make clear which confounders were adjusted for and why they were included | 8-9 |
|  |  | (b) Report category boundaries when continuous variables were categorized | N/A |
|  |  | (c) If relevant, consider translating estimates of relative risk into absolute risk for a meaningful time period | N/A |
| Other analyses | 17 | Report other analyses done—eg analyses of subgroups and interactions, and sensitivity analyses | 8-9 |
| **Discussion** |  |  |  |
| Key results | 18 | Summarise key results with reference to study objectives | 10-12 |
| Limitations | 19 | Discuss limitations of the study, taking into account sources of potential bias or imprecision. Discuss both direction and magnitude of any potential bias | 12 |
| Interpretation | 20 | Give a cautious overall interpretation of results considering objectives, limitations, multiplicity of analyses, results from similar studies, and other relevant evidence | 10-12 |
| Generalisability | 21 | Discuss the generalisability (external validity) of the study results | 10-12 |
| **Other information** |  |  |  |
| Funding | 22 | Give the source of funding and the role of the funders for the present study and, if applicable, for the original study on which the present article is based | N/A |

\*Give information separately for cases and controls in case-control studies and, if applicable, for exposed and unexposed groups in cohort and cross-sectional studies.

## Notes

### Competing Interest Statement

The authors have declared no competing interest.

### Funding Statement

None

### Author Declarations

Cleveland Clinic institutional review board

